# Protection conferred by COVID-19 vaccination, prior SARS-CoV-2 infection, or hybrid immunity against Omicron-associated severe outcomes among community-dwelling adults

**DOI:** 10.1101/2023.08.24.23294503

**Authors:** Nelson Lee, Lena Nguyen, Peter C. Austin, Kevin A. Brown, Ramandip Grewal, Sarah A Buchan, Sharifa Nasreen, Jonathan Gubbay, Kevin L Schwartz, Mina Tadrous, Kumanan Wilson, Sarah E Wilson, Jeffrey C Kwong, Canadian Immunization Research Network (CIRN) Provincial Collaborative Network (PCN) investigators

## Abstract

**Introduction:** We assessed protection conferred by COVID-19 vaccines and/or prior SARS-CoV-2 infection against Omicron-associated severe outcomes during successive sublineage-predominant periods.

**Methods:** We used a test-negative design to estimate protection by vaccines and/or prior infection against hospitalization/death among community-dwelling, PCR-tested adults aged ≥50 years in Ontario, Canada between January 2, 2022 and June 30, 2023. Multivariable logistic regression was used to estimate the relative change in the odds of hospitalization/death with each vaccine dose (2−5) and/or prior PCR-confirmed SARS-CoV-2 infection (compared with unvaccinated, uninfected subjects) up to 15 months since the last vaccination or infection.

**Results:** We included 18,526 cases with Omicron-associated severe outcomes and 90,778 test-negative controls. Vaccine protection was high during BA.1/BA.2 predominance, but was generally <50% during periods of BA.4/BA.5 and BQ/XBB predominance without boosters. A third/fourth dose transiently increased protection during BA.4/BA.5 predominance (*third-dose,* 6-month: 68%, 95%CI 63%−72%; *fourth-dose,* 6-month: 80%, 95%CI 77%−83%), but was lower and waned quickly during BQ/XBB predominance (*third-dose,* 6-month: 59%, 95%CI 48%−67%; 12-month: 49%, 95%CI 41%−56%; *fourth-dose,* 6-month: 62%, 95%CI 56%−68%, 12-months: 51%, 95%CI 41%−56%). Hybrid immunity conferred nearly 90% protection throughout BA.1/BA.2 and BA.4/BA.5 predominance, but was reduced during BQ/XBB predominance (*third-dose,* 6-month: 60%, 95%CI 36%−75%; *fourth-dose,* 6-month: 63%, 95%CI 42%−76%). Protection was restored with a fifth dose (bivalent; 6-month: 91%, 95%CI 79%−96%). Prior infection alone did not confer lasting protection.

**Conclusion:** Protection from COVID-19 vaccines and/or prior SARS-CoV-2 infections against severe outcomes is reduced when immune-evasive variants/subvariants emerge and may also wane over time. Our findings support a variant-adapted booster vaccination strategy with periodic review.

## INTRODUCTION

Omicron became the globally predominant circulating variant of SARS-CoV-2 in early 2022, and since has evolved into ever more transmissible and immune-evasive lineages and sublineages [1,2]. In the Canadian province of Ontario, the first booster (third dose) was offered to all community-dwelling adults by December 2021, and the second booster (fourth dose) to those aged ≥60 years in April 2022, and to all adults by July 2022 [3,4]. On September 1, 2022, the Moderna BA.1 bivalent vaccine was authorized by Health Canada for adults aged ≥18 years, and on October 7, 2022, the Pfizer-BioNTech BA.4/BA.5 bivalent vaccine was authorized for individuals aged ≥12 years. Moderna BA.1 and Pfizer-BioNTech BA.4/BA.5 were introduced in Ontario through a population-based fall booster program shortly after authorization, in September and October 2022, respectively [5,6]. Following their introduction, bivalent vaccines were the preferred products, however, Canada’s advisory committee on immunization recommended receiving *any* available mRNA booster dose product to ensure timely protection [7]. Therefore, monovalent mRNA vaccines were still available and accessible as booster doses. As of May 2023, over 70% of Ontario adults aged ≥50 years had received at least one booster dose of the COVID-19 vaccine [8].

While booster doses have been reported to restore some protection against Omicron [5,9,10,11], it is unclear if such protection is sustained and to what extent emergent sublineages will impact vaccine effectiveness. Vaccine effectiveness estimates of booster doses during periods when more immune-evasive sublineages, such as BA.4/BA.5, BQ, and XBB, have become predominant are lacking. Furthermore, several studies have suggested that prior infection alone or ‘hybrid immunity’ (from a combination of vaccination and a recovered SARS-CoV-2 infection) may offer sustained protection against Omicron-associated severe outcomes [12,13,14]. To inform future vaccination strategies, we investigated possible waning of protection from booster vaccine doses and/or the influence of prior SARS-CoV-2 infections against Omicron-related severe outcomes, from when Omicron first emerged and the following 18 months as the variant evolved.

## METHODS

### Study design, population, and data sources

We conducted a population-based, test-negative design study using linked databases in Ontario to investigate protection conferred by COVID-19 mRNA vaccination and/or prior SARS-CoV-2 infection against Omicron-associated severe outcomes (hospitalization or death) over time.

The study population included community-dwelling adults aged ≥50 years who underwent ≥1 polymerase chain reaction (PCR) test for SARS-CoV-2 during an 18-month period from January 2, 2022, to June 30, 2023. In Ontario, the Omicron variant and its sublineages represented nearly 100% of all positive samples since late January 2022. We excluded individuals who were immunocompromised, received a third dose earlier than November 3, 2021, received only 1 dose of mRNA vaccine, did not receive 1 dose of mRNA vaccine in the first 2 doses, and those with Delta variant infections from the analysis (**Appendix 1**). Among vaccinated subjects, 96% had received exclusively mRNA vaccines (Moderna Spikevax monovalent, Pfizer-BioNTech Comirnaty monovalent, Moderna Spikevax BA.1 bivalent, or Pfizer-BioNTech Comirnaty BA.4/BA.5 bivalent) [9].

We used provincial data from ICES (formerly the Institute for Clinical Evaluative Sciences), an independent, non-profit research institute whose legal status under Ontario’s health information privacy law allows it to collect and analyze health care and demographic data for health system evaluation and improvement. As described in our previous studies [3,4,5,9], SARS-CoV-2 laboratory testing, COVID-19 surveillance, COVID-19 vaccination, and health administrative datasets were linked using unique encoded identifiers and analyzed at ICES. The use of the data in this study is authorized under section 45 of Ontario’s Personal Health Information Protection Act and does not require review by a research ethics board.

### Definitions and sampling

We defined cases as COVID-19-associated hospitalization or death, due to, or partially due to SARS-CoV-2 infection as confirmed by PCR. Under Ontario’s provincial COVID-19 surveillance program, hospitalization data were entered only for patients who received in-hospital treatment and/or their duration of hospital stay was extended due to COVID-19 [5,9]. We excluded hospitalizations when the diagnostic specimen was collected >3 days post-admission. The controls were symptomatic but tested negative for SARS-CoV-2. Cases and controls were sampled by week of test; therefore, controls could be included in the study repeatedly. However, a control could not re-enter the study once classified as a case. This sampling strategy was used to help control for calendar time in the analysis.

We defined the index date as the date of specimen collection, hospitalization, or death, whichever was earliest. We classified subjects according to their vaccination status (vaccinated with ≥2 doses, or unvaccinated) and the number of doses received as of the index date. The primary series was defined as two monovalent vaccine doses, and boosters as third to seventh doses, either monovalent or bivalent (BA.1 and BA.4/BA.5 bivalent vaccines were available since September and October 2022, respectively) [6,9]. ‘Prior infection’ was defined as PCR-confirmed SARS-CoV-2 infection ≥60 days before the index date [15]. Results of rapid antigen tests were unavailable for study. We defined ‘time since the last immunogenic event’ with respect to the index date as the time since the most recent vaccine dose or a prior infection, whichever occurred later. Protection against hospitalization or death was reported at 3 monthly increments (to a maximum of 15 months), since the last immunogenic event, and according to Omicron sublineage predominant periods. A predominant period was defined as ≥50% of the sequenced samples being confirmed with a specific Omicron sublineage. In Ontario, the predominant sublineages were identified as BA.1/BA.2 (January 2−June 18, 2022), BA.4/BA.5 (June 19−November 26, 2022), and BQ/XBB (November 27, 2022−June 30, 2023) (**Figure 1**). The grouping was based on available evidence indicating differences in vaccine effectiveness between these sublineages due to immune evasion and other factors [1,2,5,7,10].

**Figure 1.**
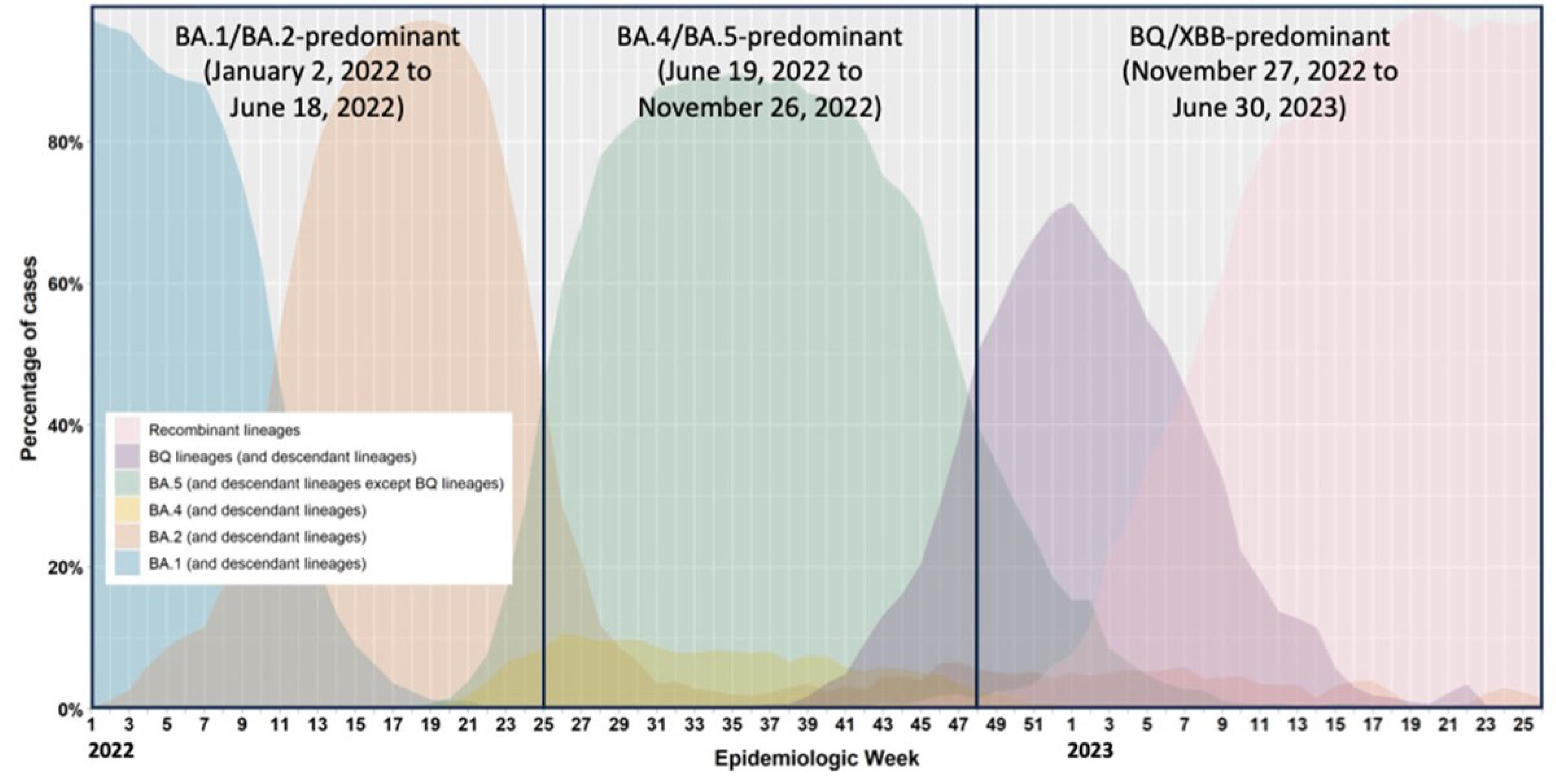
Percentage of COVID-19 cases by the most prevalent Omicron sublineages and week in Ontario, January 2, 2022 to June 30, 2023.

### Statistical analyses

We used means and proportions to describe the study population; cases and controls were compared, and the differences quantified using standardized differences (SD) during the BA.1/BA.2, BA.4/BA.5, and BQ/XBB-predominant periods (SD ≥0.1 was considered potentially clinically relevant).

We used multivariable logistic regression to estimate the relative change in the odds of severe outcomes (defined as the proportionate reduction of hospitalization or death), through vaccination and/or prior infection, compared to the reference group of subjects with neither vaccination nor documented prior infection. The estimated protection was calculated using the formula: (1-adjusted odds ratio)*100%, and reported at 3, 6, 9, 12, and 15 months since the last immunogenic event. The models were adjusted for sex, age category (50-59, 60-69, 70-79, ≥80 years), public health unit region, socio-demographic variables (neighborhood income quintile, essential worker quintile, persons per dwelling quintile, self-identified visible minority quintile), influenza vaccination status (as a proxy for health behaviors), number of SARS-CoV-2 tests within 3 months prior to December 14, 2020 (as a proxy for healthcare workers), comorbidities, receipt of home care services, and week of test (modeled using restricted cubic splines with knots at weeks 9, 28, 37, and 49) as previously described [3,4,5,9]. A 3-way interaction term between the number of doses (coded as a categorical variable), prior infection status (yes or no), and the log-transformed time since last immunogenic event was used. We further estimated the protection from prior infection in the same manner, comparing individuals with a prior infection to individuals without, at the corresponding number of vaccine doses and time since the last immunogenic event. Since the timing of vaccine dose administration with respect to a sublineage-predominant period varied, the available periods of observation for boosters could be shorter than 15 months. In the analyses where immunogenic events were restricted to vaccination, the results were interpreted as vaccine effectiveness.

We used SAS version 9.1 (SAS Institute Inc., Cary, NC) for all analyses. All tests were 2-sided, and a *P*-value <0.05 indicates statistical significance.

## RESULTS

We included 18,526 cases with Omicron-associated severe outcomes and 90,778 test-negative controls (among a total of 91,113 unique individuals), with 54,994, 29,362, and 24,948 subjects in the BA.1/BA.2-, BA.4/BA.5-, and BQ/XBB-predominant periods, respectively. In all three periods, cases were older than controls (mean ±SD, 75.9 ±11.8 *vs* 64.7 ±11.7, 78.7 ±11.2 *vs* 67.7 ±12.3, and 78.7 ±11.1 *vs* 69.8 ±12.6, respectively), and higher proportions were male (57.1% *vs* 36.9%, 55.4% *vs* 37.2%, and 56.0% *vs* 40.7%, respectively) or had comorbid conditions (91.9% *vs* 73.5%, 94.0% *vs* 77.7%, and 92.9% *vs* 80.2%, respectively); fewer had prior PCR-confirmed SARS-CoV-2 infections (0.9% *vs* 4.8%, 1.7% *vs* 6.5%, and 3.7% *vs* 6.3%, respectively), and fewer had received COVID-19 vaccines (≥2 doses, 65.9% *vs* 95.6%, 86.1% *vs* 95.2%, and 88.8% *vs* 94.4%, respectively) [**Table 1**]. In the BA.1/BA.2-predominant period, booster vaccines (3−5 doses) had been received by 2,665 (34.3%) and 35,091 (74.3%) of cases and controls, respectively. In the BA.4/BA.5-predominant period, booster vaccines (3−6 doses) had been received by 4,302 (70.1%) and 19,068 (82.1%) of cases and controls, respectively; 7.9% of these were bivalent vaccines, given as the fourth (8.1%) or fifth (97.9%) dose. In the BQ/XBB-predominant period, booster vaccines (3−7 doses) had been received by 3,459 (74.8%) and 16,571 (81.5%), respectively; 48.5% of these were bivalent vaccines (BA.1-containing, 24.1%; BA.4/BA.5-containing, 24.3%), given mostly as the fourth (51.9%) and fifth (99.2%) dose. Few subjects had received a sixth or a seventh dose. Information on bivalent booster vaccine use by period and dose count is provided in **Appendix 2**.

**Table 1:** Descriptive characteristics of community-dwelling adults tested for SARS-CoV-2 between 2-January, 2022 and 30-June, 2023 in Ontario, Canada; cases with Omicron-associated severe outcomes were compared with test-negative controls.

|  | BA.1/BA.2-predominant<br>(Jan 2, 2022 – Jun 18, 2022) |  |  | BA.4/BA.5-predominant<br>(Jun 19, 2022 – Nov 26, 2022) |  |  | BQ/XBB-predominant<br>(Nov 27, 2022 – Jun 30, 2023) |  |  |
| --- | --- | --- | --- | --- | --- | --- | --- | --- | --- |
|  | Omicron cases,<br>n (%) | Test negative<br>controls,<br>n (%)<sup>a</sup> | SD<sup>b</sup> | Omicron cases,<br>n (%) | Test negative<br>controls,<br>n (%)<sup>a</sup> | SD<sup>b</sup> | Omicron cases,<br>n (%) | Test negative<br>controls,<br>n (%)<sup>a</sup> | SD<sup>b</sup> |
|  | N=7,765 | N=47,229 |  | N=6,134 | N=23,228 |  | N=4,627 | N=20,321 |  |
| **Characteristics** |  |  |  |  |  |  |  |  |  |
| Age (mean ± standard deviation), yr | 75.87 (11.76) | 64.72 (11.67) | 0.95 | 78.73 (11.15) | 67.66 (12.27) | 0.94 | 78.66 (11.09) | 69.75 (12.64) | 0.75 |
| Male sex | 4,433 (57.1%) | 17,410 (36.9%) | 0.41 | 3,401 (55.4%) | 8,637 (37.2%) | 0.37 | 2,589 (56.0%) | 8,269 (40.7%) | 0.31 |
| Public health unit region |  |  |  |  |  |  |  |  |  |
| Central East | 466 (6.0%) | 3,572 (7.6%) | 0.06 | 455 (7.4%) | 1,499 (6.5%) | 0.04 | 351 (7.6%) | 910 (4.5%) | 0.13 |
| Central West | 1,471 (18.9%) | 7,167 (15.2%) | 0.16 | 1,169 (19.1%) | 3,803 (16.4%) | 0.19 | 925 (20.0%) | 3,872 (19.1%) | 0.17 |
| Durham | 232 (3.0%) | 2,488 (5.3%) | 0.30 | 124 (2.0%) | 631 (2.7%) | 0.48 | 71 (1.5%) | 308 (1.5%) | 0.56 |
| Eastern | 644 (8.3%) | 2,045 (4.3%) | 0.15 | 542 (8.8%) | 967 (4.2%) | 0.21 | 455 (9.8%) | 1,093 (5.4%) | 0.17 |
| North | 684 (8.8%) | 8,994 (19.0%) | 0.02 | 505 (8.2%) | 5,941 (25.6%) | 0.01 | 413 (8.9%) | 6,158 (30.3%) | 0.19 |
| Ottawa | 344 (4.4%) | 886 (1.9%) | 0.15 | 391 (6.4%) | 484 (2.1%) | 0.15 | 280 (6.1%) | 539 (2.7%) | 0.08 |
| Peel | 898 (11.6%) | 5,211 (11.0%) | 0.16 | 673 (11.0%) | 2,512 (10.8%) | 0.19 | 209 (4.5%) | 1,901 (9.4%) | 0.31 |
| South West | 986 (12.7%) | 8,463 (17.9%) | 0.12 | 851 (13.9%) | 4,536 (19.5%) | 0.23 | 727 (15.7%) | 3,804 (18.7%) | 0.35 |
| Toronto | 1,445 (18.6%) | 6,107 (12.9%) | 0.02 | 941 (15.3%) | 2,134 (9.2%) | 0.03 | 770 (16.6%) | 1,385 (6.8%) | 0.01 |
| York | 563 (7.3%) | 2,150 (4.6%) | 0.10 | 467 (7.6%) | 621 (2.7%) | 0.07 | 411 (8.9%) | 275 (1.4%) | 0.02 |
| Missing | 32 (0.4%) | 146 (0.3%) | 0.12 | 16 (0.3%) | 100 (0.4%) | 0.05 | 15 (0.3%) | 76 (0.4%) | 0.00 |
| Household income quintile |  |  |  |  |  |  |  |  |  |
| 1 (lowest) | 2,198 (28.3%) | 9,609 (20.3%) | 0.19 | 1,490 (24.3%) | 5,236 (22.5%) | 0.04 | 1,188 (25.7%) | 4,878 (24.0%) | 0.04 |
| 2 | 1,731 (22.3%) | 9,343 (19.8%) | 0.06 | 1,332 (21.7%) | 4,961 (21.4%) | 0.01 | 1,000 (21.6%) | 4,514 (22.2%) | 0.02 |
| 3 | 1,447 (18.6%) | 9,155 (19.4%) | 0.02 | 1,234 (20.1%) | 4,323 (18.6%) | 0.04 | 867 (18.7%) | 3,855 (19.0%) | 0.01 |
|  | Omicron cases,<br>n (%) | Test negative<br>controls,<br>n (%)<sup>a</sup> | SD<sup>b</sup> | Omicron cases,<br>n (%) | Test negative<br>controls,<br>n (%)<sup>a</sup> | SD<sup>b</sup> | Omicron cases,<br>n (%) | Test negative<br>controls,<br>n (%)<sup>a</sup> | SD<sup>b</sup> |
| 4 | 1,285 (16.5%) | 9,249 (19.6%) | 0.08 | 1,097 (17.9%) | 4,351 (18.7%) | 0.02 | 843 (18.2%) | 3,583 (17.6%) | 0.02 |
| 5 (highest) | 1,076 (13.9%) | 9,724 (20.6%) | 0.18 | 956 (15.6%) | 4,267 (18.4%) | 0.07 | 716 (15.5%) | 3,426 (16.9%) | 0.04 |
| Missing | 28 (0.4%) | 149 (0.3%) | 0.01 | 25 (0.4%) | 90 (0.4%) | 0.00 | 13 (0.3%) | 65 (0.3%) | 0.01 |
| Essential workers quintile |  |  |  |  |  |  |  |  |  |
| 1 (0–32.5) | 1,015 (13.1%) | 7,449 (15.8%) | 0.08 | 989 (16.1%) | 3,125 (13.5%) | 0.08 | 773 (16.7%) | 2,485 (12.2%) | 0.13 |
| 2 (32.5–42.3) | 1,539 (19.8%) | 10,346 (21.9%) | 0.05 | 1,210 (19.7%) | 4,853 (20.9%) | 0.03 | 930 (20.1%) | 4,203 (20.7%) | 0.01 |
| 3 (42.3–49.8) | 1,630 (21.0%) | 10,334 (21.9%) | 0.02 | 1,322 (21.6%) | 5,250 (22.6%) | 0.03 | 894 (19.3%) | 4,551 (22.4%) | 0.08 |
| 4 (50.0–57.5) | 1,749 (22.5%) | 9,639 (20.4%) | 0.05 | 1,306 (21.3%) | 4,938 (21.3%) | 0.00 | 1,002 (21.7%) | 4,511 (22.2%) | 0.01 |
| 5 (57.5–100) | 1,777 (22.9%) | 9,092 (19.3%) | 0.09 | 1,279 (20.9%) | 4,876 (21.0%) | 0.00 | 1,007 (21.8%) | 4,428 (21.8%) | 0.00 |
| Missing | 55 (0.7%) | 369 (0.8%) | 0.01 | 28 (0.5%) | 186 (0.8%) | 0.04 | 21 (0.5%) | 143 (0.7%) | 0.03 |
| Persons per dwelling quintile |  |  |  |  |  |  |  |  |  |
| 1 (0–2.1) | 60 (0.8%) | 390 (0.8%) | 0.01 | 34 (0.6%) | 202 (0.9%) | 0.04 | 22 (0.5%) | 167 (0.8%) | 0.04 |
| 2 (2.2–2.4) | 2,031 (26.2%) | 10,599 (22.4%) | 0.09 | 1,686 (27.5%) | 5,938 (25.6%) | 0.04 | 1,335 (28.9%) | 5,792 (28.5%) | 0.01 |
| 3 (2.5–2.6) | 1,540 (19.8%) | 10,368 (22.0%) | 0.05 | 1,255 (20.5%) | 5,454 (23.5%) | 0.07 | 993 (21.5%) | 5,206 (25.6%) | 0.10 |
| 4 (2.7–3.0) | 1,088 (14.0%) | 6,385 (13.5%) | 0.01 | 833 (13.6%) | 3,283 (14.1%) | 0.02 | 633 (13.7%) | 2,863 (14.1%) | 0.01 |
| 5 (3.1–5.7) | 1,532 (19.7%) | 10,050 (21.3%) | 0.04 | 1,226 (20.0%) | 4,540 (19.5%) | 0.01 | 969 (20.9%) | 3,631 (17.9%) | 0.08 |
| Missing | 1,514 (19.5%) | 9,437 (20.0%) | 0.01 | 1,100 (17.9%) | 3,811 (16.4%) | 0.04 | 675 (14.6%) | 2,662 (13.1%) | 0.04 |
| Self-identified visible minority<br>quintile |  |  |  |  |  |  |  |  |  |
| 1 (0.0–2.2) | 1,484 (19.1%) | 10,871 (23.0%) | 0.10 | 1,199 (19.5%) | 6,156 (26.5%) | 0.17 | 960 (20.7%) | 5,908 (29.1%) | 0.19 |
| 2 (2.2–7.5) | 1,333 (17.2%) | 10,501 (22.2%) | 0.13 | 1,168 (19.0%) | 5,487 (23.6%) | 0.11 | 981 (21.2%) | 5,190 (25.5%) | 0.10 |
| 3 (7.5–18.7) | 1,364 (17.6%) | 8,459 (17.9%) | 0.01 | 1,220 (19.9%) | 4,148 (17.9%) | 0.05 | 930 (20.1%) | 3,554 (17.5%) | 0.07 |
| 4 (18.7–43.5) | 1,546 (19.9%) | 8,104 (17.2%) | 0.07 | 1,169 (19.1%) | 3,505 (15.1%) | 0.11 | 857 (18.5%) | 2,949 (14.5%) | 0.11 |
| 5 (43.5–100) | 1,984 (25.6%) | 8,925 (18.9%) | 0.16 | 1,350 (22.0%) | 3,748 (16.1%) | 0.15 | 878 (19.0%) | 2,577 (12.7%) | 0.17 |
|  | Omicron cases,<br>n (%) | Test negative<br>controls,<br>n (%)<sup>a</sup> | SD<sup>b</sup> | Omicron cases,<br>n (%) | Test negative<br>controls,<br>n (%)<sup>a</sup> | SD<sup>b</sup> | Omicron cases,<br>n (%) | Test negative<br>controls,<br>n (%)<sup>a</sup> | SD<sup>b</sup> |
| Missing | 54 (0.7%) | 369 (0.8%) | 0.01 | 28 (0.5%) | 184 (0.8%) | 0.04 | 21 (0.5%) | 143 (0.7%) | 0.03 |
| Receipt of 2019-2020 and/or<br>2020-2021 influenza<br>vaccination (2019-2021) | 3,838 (49.4%) | 24,342 (51.5%) | 0.04 | 3,905 (63.7%) | 12,654 (54.5%) | 0.19 | 2,991 (64.6%) | 11,216<br>(55.2%) | 0.19 |
| No. of SARS-CoV-2 tests prior<br>to December 14, 2020 |  |  |  |  |  |  |  |  |  |
| 0 | 6,859 (88.3%) | 34,063 (72.1%) | 0.42 | 5,429 (88.5%) | 17,254 (74.3%) | 0.37 | 4,103 (88.7%) | 15,830<br>(77.9%) | 0.29 |
| 1 | 630 (8.1%) | 7,045 (14.9%) | 0.21 | 491 (8.0%) | 2,902 (12.5%) | 0.15 | 361 (7.8%) | 2,176 (10.7%) | 0.10 |
| ≥2 | 276 (3.6%) | 6,121 (13.0%) | 0.35 | 214 (3.5%) | 3,072 (13.2%) | 0.36 | 163 (3.5%) | 2,315 (11.4%) | 0.30 |
| Any comorbidity (other than<br>immunocompromised) | 7,133 (91.9%) | 34,709 (73.5%) | 0.50 | 5,769 (94.0%) | 18,047 (77.7%) | 0.48 | 4,300 (92.9%) | 16,295<br>(80.2%) | 0.38 |
| Receipt of home care services |  |  |  |  |  |  |  |  |  |
| None | 7,064 (91.0%) | 45,178 (95.7%) | 0.19 | 5,386 (87.8%) | 21,610 (93.0%) | 0.18 | 4,200 (90.8%) | 18,898<br>(93.0%) | 0.08 |
| Short stay | 250 (3.2%) | 1,072 (2.3%) | 0.06 | 275 (4.5%) | 855 (3.7%) | 0.04 | 169 (3.7%) | 754 (3.7%) | 0.00 |
| Long stay | 419 (5.4%) | 891 (1.9%) | 0.19 | 437 (7.1%) | 678 (2.9%) | 0.19 | 245 (5.3%) | 609 (3.0%) | 0.12 |
| Palliative | 32 (0.4%) | 88 (0.2%) | 0.04 | 36 (0.6%) | 85 (0.4%) | 0.03 | 13 (0.3%) | 60 (0.3%) | 0.00 |
| **Most recent vaccine dose** |  |  |  |  |  |  |  |  |  |
| Unvaccinated | 2,650 (34.1%) | 2,074 (4.4%) | 0.81 | 850 (13.9%) | 1,109 (4.8%) | 0.32 | 516 (11.2%) | 1,139 (5.6%) | 0.20 |
| Second | 2,450 (31.6%) | 10,064 (21.3%) | 0.23 | 982 (16.0%) | 3,051 (13.1%) | 0.08 | 652 (14.1%) | 2,611 (12.8%) | 0.04 |
| Third | 2,481 (32.0%) | 32,996 (69.9%) | 0.82 | 2,430 (39.6%) | 10,784 (46.4%) | 0.14 | 1,342 (29.0%) | 6,043 (29.7%) | 0.02 |
| Fourth | 184 (2.4%) | 2090-2094<sup>c</sup><br>(4.4%) | 0.11 | 1,702 (27.7%) | 7,349 (31.6%) | 0.09 | 1,074 (23.2%) | 5,415 (26.6%) | 0.08 |
| Fifth | 0 (0.0%) | ≤5<sup>c</sup><br>(≤0.01%) | 0.01 | 165-169<sup>c</sup><br>(2.7-2.8%) | 930-934<sup>c</sup><br>(4.0%) | 0.07 | 1,011 (21.9%) | 4,892 (24.1%) | 0.05 |
|  | Omicron cases,<br>n (%) | Test negative<br>controls,<br>n (%)<sup>a</sup> | SD<sup>b</sup> | Omicron cases,<br>n (%) | Test negative<br>controls,<br>n (%)<sup>a</sup> | SD<sup>b</sup> | Omicron cases,<br>n (%) | Test negative<br>controls,<br>n (%)<sup>a</sup> | SD<sup>b</sup> |
| Sixth | - | - | - | ≤5<sup>c</sup><br>(≤0.1%) | ≤5<sup>c</sup><br>(≤0.02%) | 0.01 | 32 (0.7%) | 216-220<sup>c</sup><br>(1.1%) | 0.04 |
| Seventh | - | - | - | - | - | - | 0 (0.0%) | ≤5<sup>c</sup><br>(≤0.02%) | 0.02 |
| **Prior infection** (positive SARS-CoV-2 PCR test, >60 days prior) | 71 (0.9%) | 2,267 (4.8%) | 0.24 | 105 (1.7%) | 1,509 (6.5%) | 0.24 | 169 (3.7%) | 1,288 (6.3%) | 0.12 |

<sup>a</sup> Proportion reported, unless stated otherwise.
<sup>b</sup> SD = standardized difference. Standardized differences of >0.10 are considered clinically relevant. Cases with Omicron-associated severe outcomes were compared with test-negative controls in each omicron sublineage-predominant period.
<sup>c</sup> Due to institutional privacy policies, any cells ≤5 (except for missing values) must be suppressed and ranges must be provided for complementary cells to prevent back calculation. Other demographic descriptives of the Ontario cohort had been reported [5,9].

### Vaccine protection against severe outcomes

Among subjects with no PCR-confirmed prior infection (>94% of this cohort), we estimated protection (i.e., vaccine effectiveness) at 3, 6, 9, 12, and 15 months since the last immunogenic event (i.e., last vaccine dose), where applicable [**Figure 2**, panel left (grey)]. With the primary 2-dose series, protection against severe outcomes throughout the BA.1/BA.2-predominant period was approximately 80% (e.g., at 6 months: 80%; 95%CI, 78%−82%). During the BA.4/BA.5-predominant period, protection declined markedly to below 50% (e.g., at 6 months: 46%; 95%CI, 33%−57%), and further reduced to below 30% during the BQ/XBB-predominant period. Booster doses increased the levels of protection during the BA.1/BA.2-predominant period (e.g., *third-dose* at 6 months: 94%; 95%CI, 93%−95), but to a much lesser extent during the BA.4/BA.5-predominant period; although protection tended to increase with more doses (e.g., *third-dose* at 6 months: 68%; 95%CI, 63%−72%; *fourth-dose* at 6 months: 80%; 95%CI, 77%−83%). During the BQ/XBB-predominant period, estimates of boosters’ protection against severe outcomes were further reduced, and waned more quickly over time (e.g., *third-dose* at 6 months: 59%; 95%CI, 48%−67%; 12 months: 49%; 95%CI, 41%−56%; *fourth-dose* at 6 months: 62%; 95%CI, 56%−68%; 12 months: 51%; 95%CI, 41%−56%; *fifth-dose* at 6 months: 69%; 95%CI, 63%−74%).

**Figure 2.**
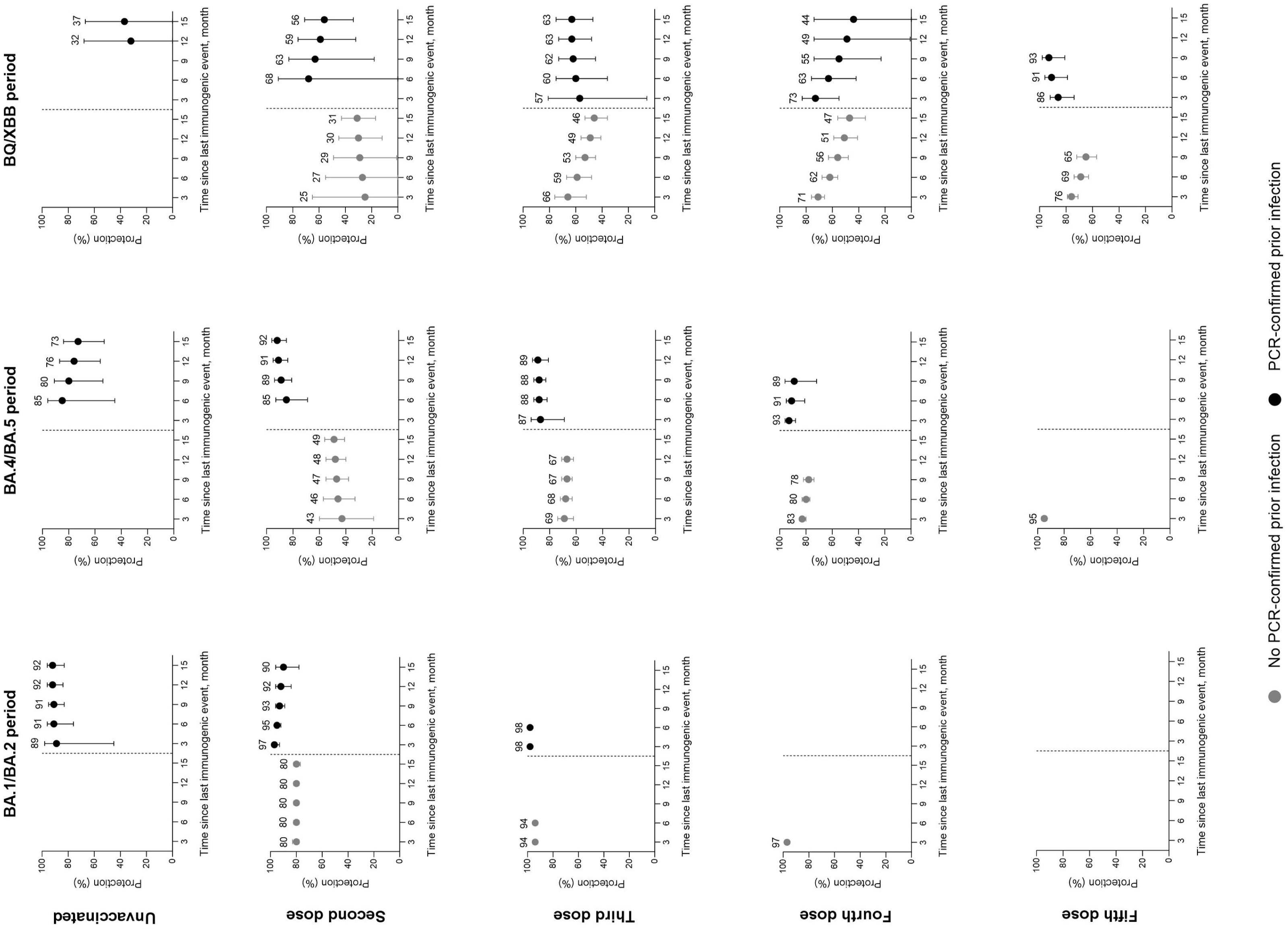
Estimated protection (proportionate reduction) against severe outcomes and 95% confidence intervals by time since the last immunogenic event, according to prior infection status. Results were reported for each sublineage (BA.1/BA.2-, BA.4/BA.5, BQ/XBB) predominant period.

### Protection conferred by prior SARS-CoV-2 infection

In the population subset with documented, PCR-confirmed prior infections, and who were vaccinated, we estimated protection conferred by hybrid immunity against severe outcomes at 3, 6, 9, 12, and 15 months since the last immunogenic event, where applicable [**Figure 2**, panel right (black)]. We observed high levels of protection (>90%) during the BA.1/BA.2-predominant period, lasting for at least 12 months. Protection was slightly lower during the BA.4/BA.5-predominant period but increased with more booster doses (e.g., *third-dose* at 6 months 88%; 95%CI, 82%−92%; *fourth-dose* at 6 months 91%; 95%CI, 81%−95%), and remained high for 9−12 months. During the BQ/XBB-predominant period, however, protection was reduced for those who received only the primary series, and for third and fourth dose recipients; and it tended to wane more quickly over time (e.g., *third-dose* at 6 months: 60%; 95%CI, 36%−75%, 12 months 63%; 95%CI, 48%−73%; *fourth-dose* at 6 months: 63%; 95%CI, 42%−76%, 12 months 49%; 95%CI, 1%−47%). Protection was restored for those who had received a fifth dose (at 6 months: 91%; 95%CI, 79%−96%).

Among vaccinated subjects, prior infection (compared with no prior infection) was associated with significantly greater protection against severe outcomes for the corresponding number of vaccine doses and time points, during the BA.1/BA.2-predominant (for *second*-dose, at 3−12 months; *third-*dose, at 3−6 months) and BA.4/BA.5-predominant (for *second*-dose, at 6−15 months; *third*-dose, at 3−12 months; *fourth*-dose, at 3−6 months) periods (**Table 2**). It was generally not significant at most time points during the BQ/XBB-predominant period, except for those who received a fifth dose, at 6−9 months.

**Table 2.**
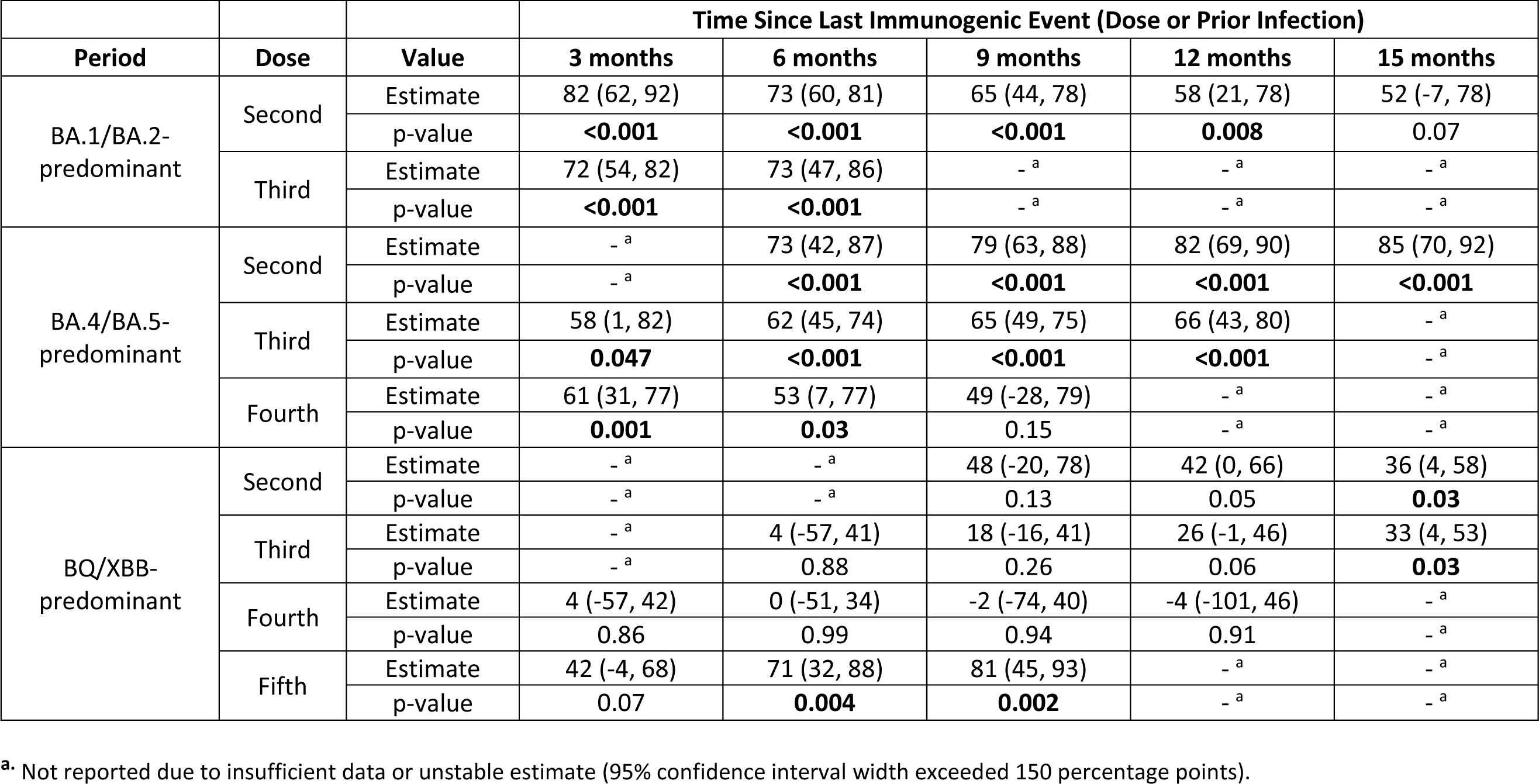
Marginal protection against Omicron-associated severe outcomes from documented prior SARS-CoV-2 infection among vaccinated individuals, stratified by period of Omicron sublineage predominance, number of COVID-19 vaccine doses received, and time since last immunogenic event (either COVID-19 vaccine dose or prior SARS-CoV-2 infection)

| Period | Dose | Value | Time Since Last Immunogenic Event (Dose or Prior Infection) |  |  |  |  |
| --- | --- | --- | --- | --- | --- | --- | --- |
|  |  |  | 3 months | 6 months | 9 months | 12 months | 15 months |
| BA.1/BA.2-predominant | Second | Estimate | 82 (62, 92) | 73 (60, 81) | 65 (44, 78) | 58 (21, 78) | 52 (-7, 78) |
|  |  | p-value | **<0.001** | **<0.001** | **<0.001** | **0.008** | 0.07 |
|  | Third | Estimate | 72 (54, 82) | 73 (47, 86) | -<sup>a</sup> | -<sup>a</sup> | -<sup>a</sup> |
|  |  | p-value | **<0.001** | **<0.001** | -<sup>a</sup> | -<sup>a</sup> | -<sup>a</sup> |
| BA.4/BA.5-predominant | Second | Estimate | -<sup>a</sup> | 73 (42, 87) | 79 (63, 88) | 82 (69, 90) | 85 (70, 92) |
|  |  | p-value | -<sup>a</sup> | **<0.001** | **<0.001** | **<0.001** | **<0.001** |
|  | Third | Estimate | 58 (1, 82) | 62 (45, 74) | 65 (49, 75) | 66 (43, 80) | -<sup>a</sup> |
|  |  | p-value | **0.047** | **<0.001** | **<0.001** | **<0.001** | -<sup>a</sup> |
|  | Fourth | Estimate | 61 (31, 77) | 53 (7, 77) | 49 (-28, 79) | -<sup>a</sup> | -<sup>a</sup> |
|  |  | p-value | **0.001** | **0.03** | 0.15 | -<sup>a</sup> | -<sup>a</sup> |
| BQ/XBB-predominant | Second | Estimate | -<sup>a</sup> | -<sup>a</sup> | 48 (-20, 78) | 42 (0, 66) | 36 (4, 58) |
|  |  | p-value | -<sup>a</sup> | -<sup>a</sup> | 0.13 | 0.05 | **0.03** |
|  | Third | Estimate | -<sup>a</sup> | 4 (-57, 41) | 18 (-16, 41) | 26 (-1, 46) | 33 (4, 53) |
|  |  | p-value | -<sup>a</sup> | 0.88 | 0.26 | 0.06 | **0.03** |
|  | Fourth | Estimate | 4 (-57, 42) | 0 (-51, 34) | -2 (-74, 40) | -4 (-101, 46) | -<sup>a</sup> |
|  |  | p-value | 0.86 | 0.99 | 0.94 | 0.91 | -<sup>a</sup> |
|  | Fifth | Estimate | 42 (-4, 68) | 71 (32, 88) | 81 (45, 93) | -<sup>a</sup> | -<sup>a</sup> |
|  |  | p-value | 0.07 | **0.004** | **0.002** | -<sup>a</sup> | -<sup>a</sup> |

<sup>a</sup>. Not reported due to insufficient data or unstable estimate (95% confidence interval width exceeded 150 percentage points).

Among the unvaccinated, prior infection alone was associated with protection generally >90% throughout the BA.1/BA.2-predominant period. However, during the BA.4/BA.5-predominant period, protection waned from 85% at 6 months to 76% at 12 months, and was only 32% at 12 months during the BQ/XBB-predominant period.

## DISCUSSION

In this population-based study spanning three Omicron sublineage-predominant periods over 1.5 years, we found that vaccine effectiveness against severe outcomes of the two-dose primary vaccination series alone was generally below 50% during the BA.4/BA.5- and BQ/XBB-predominant periods. Booster vaccine doses transiently increased protection during the BA.4/BA.5-predominant period, but protection was lower and waned relatively quickly over months during the BQ/XBB-predominant period. Prior infection alone also did not provide lasting protection. Hybrid immunity was associated with higher levels of protection in earlier periods but was reduced and waned more quickly during the BQ/XBB-predominant period. Our results suggest that a variant-adapted booster vaccination strategy is necessary as the virus continues to evolve.

Our findings are in line with a recent meta-analysis that found marked reduction of vaccine effectiveness with the primary series against hospitalizations and deaths in the Omicron era; booster vaccine doses partially restored protection but it waned over months [11]. This study provides new data on Omicron sublineages and extended observation up to 15 months since the last vaccine dose. Our point estimates of protection conferred by the primary series among subjects without documented prior infection were all <50% during the BA.4/BA.5-predominant and <30% during the BQ/XBB-predominant periods; even with a booster (third or fourth) dose, protection was generally <80% during the former and <62% after 6 months during the latter period, and declined progressively as time elapsed. We found that repeated booster dosing, alongside with the introduction of bivalent vaccines, given mostly as a fifth dose, restored protection to some degree (about 70% at 6 months), similar to other studies [16,17,18,19,20,21,22]. Vaccine effectiveness of different bivalent vaccine products among adults in Ontario has been explored separately [9].

On hybrid immunity, a recent meta-regression analysis of 15 studies showed high levels of protection (>90%) against severe outcomes with either primary series or booster vaccine doses, which lasted for at least 6 months [12]. However, the analysis only included studies before June 2022, thus the impact of Omicron sublineages that emerged later (i.e., BA.4, BA.5, BQ, and XBB) were not examined. Our study found similar point estimates at around 90%, lasting for >6 months, during both BA.1/BA.2- and BA.4/BA.5-predominant periods; and we showed significantly greater protection with hybrid immunity compared with vaccination alone. Notably, such protection dropped significantly to generally <70% when the more immune-evasive BQ/XBB sublineages became predominant, only to be restored with a fifth dose (nearly all being bivalent vaccines, **Appendix 2**)[23,24]. While earlier studies had reported nearly 80% protection against severe outcomes with prior infection alone (i.e., unvaccinated), lasting for about 6 months in the BA.1/BA.2- and BA.4/BA.5-predominant periods [12,13,14], we observed progressive decline from 6 to 15 months during the latter period. Protection was very low during the BQ/XBB-predominant period. Taken together, we found that neither vaccine-induced, infection-induced, nor hybrid immunity provide long-lasting protection against severe COVID-19 outcomes as virus sublineages continue to evolve over time. Our data support the approach to review vaccine composition based on continual surveillance and offer booster doses periodically, especially for high-risk individuals, even in immune-experienced populations where vaccine coverage is high and prior SARS-CoV-2 infection is prevalent [25].

The strengths of our study include being population-based, the large sample size, evaluation of multiple Omicron sublineage-predominant periods, stratification by the latest vaccine dose and longer observation durations, and adjustment for available confounding factors. Our study has several limitations. We could only capture prior infections confirmed by PCR tests performed in healthcare settings in Ontario; results of rapid antigen tests were unavailable.

Canada-wide data indicate that infection-acquired seropositivity has risen steadily during the Omicron waves among older adults (it approached 62% by February 2023) [26], thus undocumented infections are probable. Despite some level of infection-induced immunity likely existing among vaccinated subjects and the unvaccinated reference group, we still found greater risk reduction with prior PCR-confirmed disease, suggesting that the contemporary observed vaccine effectiveness has been modified by accumulative, collective immune experience in the population. It is unclear if pauci-symptomatic or asymptomatic infections (untested or diagnosed by antigen tests in community settings) generate less durable immunity, which deserves further study [27,28]. Secondly, we were unable to evaluate vaccine effectiveness against specific Omicron sublineage due to limited sequencing data, but our categorization according to sublineage-predominant periods allowed study of the temporal changes as the virus evolved. Lastly, we acknowledge that at the time of writing, the observation periods for the fifth doses were relatively short, and few subjects had received the sixth/seventh doses.

In conclusion, among community-dwelling adults, protection from COVID-19 vaccines and/or prior SARS-CoV-2 infections against severe outcomes is reduced when immune-evasive variants/subvariants emerge and may also wane over time. Protection was overall enhanced with hybrid immunity, whereas neither prior infection nor prior vaccination alone provides lasting protection. Our results indicate that a variant-adapted booster vaccination strategy with periodic review is likely necessary even in immune-experienced populations with high vaccine coverage and/or prior infection prevalence.

## Funding

This work was supported by funding from the Canadian Immunization Research Network (CIRN) through a grant from the Public Health Agency of Canada and the Canadian Institutes of Health Research (CNF 151944), and also by funding from the Public Health Agency of Canada, through the Vaccine Surveillance Working Party and the COVID-19 Immunity Task Force. This study was supported by Public Health Ontario and by ICES, which is funded by an annual grant from the Ontario Ministry of Health (MOH) and Ministry of Long-Term Care (MLTC). This work was also supported by the Ontario Health Data Platform (OHDP), a Province of Ontario initiative to support Ontario’s ongoing response to COVID-19 and its related impacts. Jeffrey C. Kwong is supported by a Clinician-Scientist Award from the University of Toronto Department of Family and Community Medicine. The study sponsors did not participate in the design and conduct of the study; collection, management, analysis and interpretation of the data; preparation, review or approval of the manuscript; or the decision to submit the manuscript for publication.

## Data Availability

All data produced in the present work are contained in the manuscript

## Acknowledgements and Disclaimers

We would like to acknowledge the Canadian Immunization Research Network (CIRN) Provincial Collaborative Network (PCN) Investigators, Public Health Ontario for access to vaccination data from COVaxON, case-level data from the Public Health Case and Contact Management Solution (CCM) and COVID-19 laboratory data, as well as assistance with data interpretation. We also thank the staff of Ontario’s public health units who are responsible for COVID-19 case and contact management and data collection within CCM. We thank IQVIA Solutions Canada Inc. for use of their Drug Information File. The authors are grateful to the Ontario residents without whom this research would be impossible.

This document used data adapted from the Statistics Canada Postal Code^OM^ Conversion File, which is based on data licensed from Canada Post Corporation, and/or data adapted from the Ontario Ministry of Health Postal Code Conversion File, which contains data copied under license from ^©^Canada Post Corporation and Statistics Canada. Parts of this material are based on data and/or information compiled and provided by: MOH, Ontario Health, the Canadian Institute for Health Information, Statistics Canada, and IQVIA Solutions Canada Inc. The analyses, conclusions, opinions and statements expressed herein are solely those of the authors and do not reflect those of the funding or data sources; no endorsement is intended or should be inferred. Adapted from Statistics Canada, Canadian Census 2016. This does not constitute an endorsement by Statistics Canada of this product.

## Competing interests

Nelson Lee has previously received honoraria for consultancy work, speaking in educational programs, and/or travel support from: Shionogi Inc., Gilead Sciences Canada Inc., Janssen Inc., GlaxoSmithKline plc., Sanofi Pasteur Ltd., F. Hoffmann-La Roche Ltd., Genentech Inc., CIDARA Therapeutics Inc., Clarion Healthcare, bioStrategies, Technospert, Aligos; all <u>unrelated</u> to this work. Kumanan Wilson is a shareholder and board member of CANImmunize Inc. and has served on independent scientific advisory boards for Medicago and Moderna.

## Appendix 1

**Community-dwelling adults aged ≥50 years included in the analysis between January 2, 2022 and June 30, 2023, in Ontario, Canada**

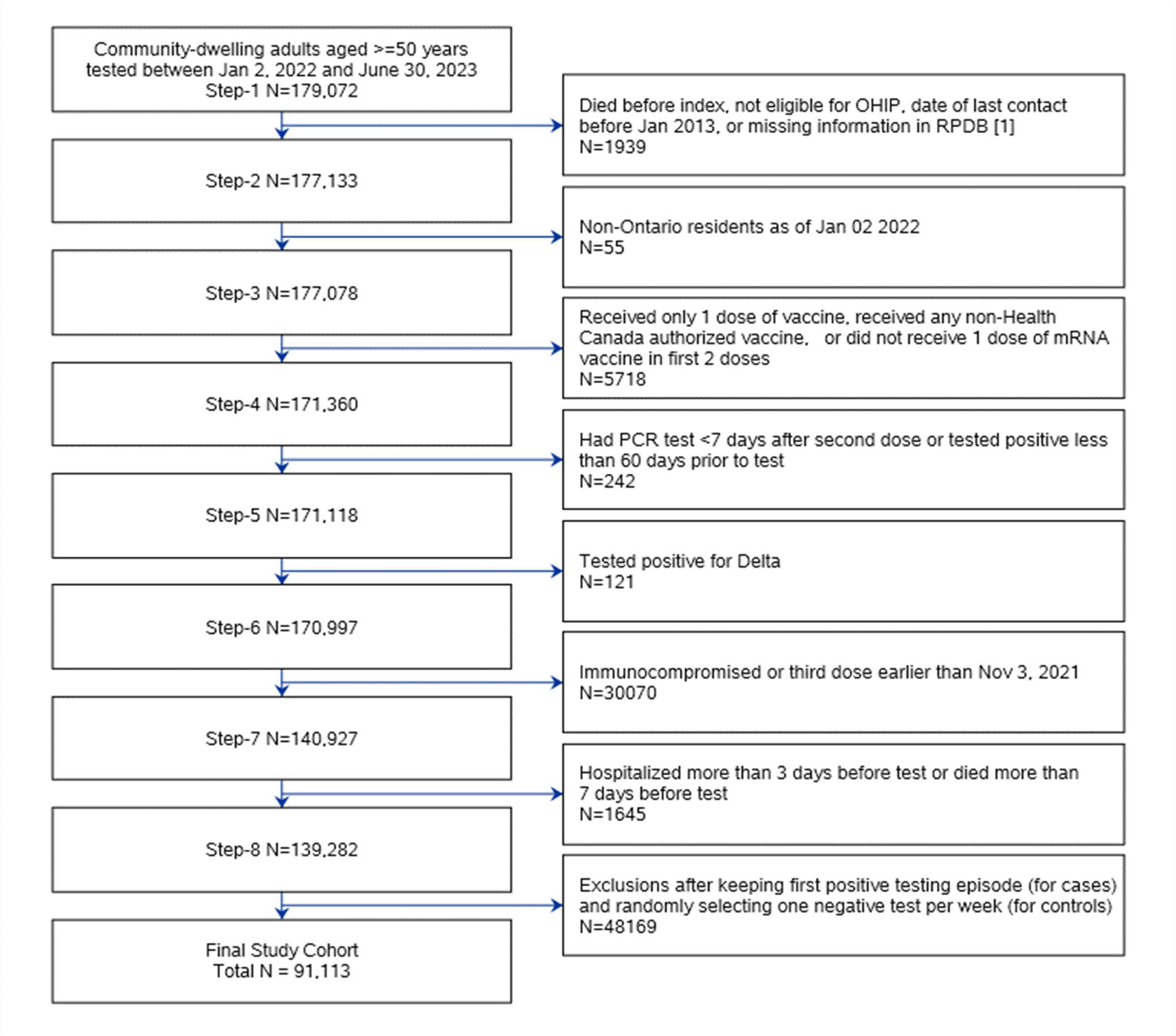

[1] OHIP: Ontaria Health Insurance Plan. RPDB: Registered Persons Database

## Appendix 2

**Bivalent Omicron-containing booster vaccine (Moderna Spikevax BA.1 or Pfizer-BioNTech Comirnaty BA.4/BA.5) use in successive variant sublineage-predominant periods.**

|  | **BA.1/BA.2-predominant**<br>(Jan 2, 2022– Jun 18, 2022)<br>n (%) | **BA.4/BA.5- predominant**<br>(Jun 19, 2022–Nov 26, 2022)<br>n (%) | **BQ/XBB-predominant**<br>(Nov 27, 2022 – June 30,<br>2023) n (%) |
| --- | --- | --- | --- |
| **most recent**<br>**vaccine dose** |  |  |  |
| *third* | 0 (0.0%) | 30 (0.2%) | 234 (3.2%) |
| *fourth* | 0 (0.0%) | 737 (8.1%) | 3,367 (51.9%) |
| *fifth* | 0 (0.0%) | 1,080 (97.9%) | 5,854 (99.2%) |
| *sixth* | 0 (0.0%) | ≤5 (100.0%) | 248 (99.2%) |

## Notes

### Author Declarations

We used provincial data from ICES (formerly the Institute for Clinical Evaluative Sciences), an independent, non-profit research institute whose legal status under Ontario's health information privacy law allows it to collect and analyze health care and demographic data for health system evaluation and improvement. As described in our previous studies [reference 3,4,5,9], SARS-CoV-2 laboratory testing, COVID-19 surveillance, COVID-19 vaccination, and health administrative datasets were linked using unique encoded identifiers and analyzed at ICES. The use of the data in this study is authorized under section 45 of Ontario's Personal Health Information Protection Act and does not require review by a research ethics board. ICES is a prescribed entity under Ontario's Personal Health Information Protection Act (PHIPA). Section 45 of PHIPA authorizes ICES to collect personal health information, without consent, for the purpose of analysis or compiling statistical information with respect to the management of, evaluation or monitoring of, the allocation of resources to or planning for all or part of the health system. Projects that use data collected by ICES under section 45 of PHIPA, and use no other data, are exempt from REB review. The use of the data in this project is authorized under section 45 and approved by ICES' Privacy and Legal Office.

## References

1. Feikin DR, Higdon MM, Andrews N, Collie S, Deloria Knoll M, Kwong JC, Link-Gelles R, Pilishvili T, Patel MK. Assessing COVID-19 vaccine effectiveness against Omicron subvariants: Report from a meeting of the World Health Organization. Vaccine. 2023 Mar 31;41(14):2329–2338.

2. UK Health and Security Agency. COVID-19 vaccine monthly surveillance reports (week 39 2021 to week 14 2023). https://www.gov.uk/government/publications/covid-19-vaccine-weekly-surveillance-reports

3. Buchan SA, Chung H, Brown KA, Austin PC, Fell DB, Gubbay JB, Nasreen S, Schwartz KL, Sundaram ME, Tadrous M, Wilson K, Wilson SE, Kwong JC. Estimated effectiveness of COVID-19 vaccines against Omicron or Delta symptomatic infection and severe outcomes. JAMA Network Open. 2022 Sep 1;5(9):e2232760.

4. Grewal R, Kitchen SA, Nguyen L, Buchan SA, Wilson SE, Costa AP, Kwong JC. Effectiveness of a fourth dose of covid-19 mRNA vaccine against the omicron variant among long term care residents in Ontario, Canada: test negative design study. BMJ. 2022 Jul 6;378:e071502.

5. Grewal R, Nguyen L, Buchan SA, Wilson SE, Nasreen S, Austin PC, Brown KA, Fell DB, Gubbay JB, Schwartz KL, Tadrous M, Wilson K, Kwong JC. Effectiveness of mRNA COVID-19 vaccine booster doses against Omicron severe outcomes. Nat Commun. 2023 Mar 7;14(1):1273.

6. Government of Ontario. Ontarians aged 18+ eligible for bivalent COVID-19 booster dose. https://news.ontario.ca/en/release/1002277/ontarians-aged-18-eligible-for-bivalent-covid-19-booster-dose

7. National Advisory Committee on Immunization (NACI). An Advisory Committee Statement (ACS). Recommendations on the use of bivalent Omicron-containing mRNA COVID-19 vaccines https://www.canada.ca/content/dam/phac-aspc/documents/services/immunization/national-advisory-committee-on-immunization-naci/recommendations-use-bivalent-Omicron-containing-mrna-covid-19-vaccines.pdf

8. Government of Canada. COVID-19 vaccination: Vaccination coverage. https://health-infobase.canada.ca/covid-19/vaccination-coverage/(last accessed 12-July, 2023)

9. Grewal R, Buchan SA, Nguyen L, Nasreen S, Austin PC, Brown KA, Fell DB, Gubbay JB, Lee N, Schwartz KL, Tadrous M, Wilson K, Wilson SE, Kwong JC; the Canadian Immunization Research Network (CIRN) Provincial Collaborative Network investigators. Effectiveness of mRNA COVID-19 monovalent and bivalent vaccine booster doses against Omicron severe outcomes among adults aged ≥50 years in Ontario, Canada. doi: 10.1101/2023.04.11.23288403 [preprint]

10. Tseng HF, Ackerson BK, Bruxvoort KJ, Sy LS, Tubert JE, Lee GS, Ku JH, Florea A, Luo Y, Qiu S, Choi SK, Takhar HS, Aragones M, Paila YD, Chavers S, Talarico CA, Qian L. Effectiveness of mRNA-1273 vaccination against SARS-CoV-2 omicron subvariants BA.1, BA.2, BA.2.12.1, BA.4, and BA.5. Nat Commun. 2023 Jan 12;14(1):189.

11. Wu N, Joyal-Desmarais K, Ribeiro PAB, Vieira AM, Stojanovic J, Sanuade C, Yip D, Bacon SL. Long-term effectiveness of COVID-19 vaccines against infections, hospitalisations, and mortality in adults: findings from a rapid living systematic evidence synthesis and meta-analysis up to December, 2022. Lancet Respir Med. 2023 Feb 10.S2213-2600(23)00015-2.

12. Bobrovitz N, Ware H, Ma X, Li Z, Hosseini R, Cao C, Selemon A, Whelan M, Premji Z, Issa H, Cheng B, Abu Raddad LJ, Buckeridge DL, Van Kerkhove MD, Piechotta V, Higdon MM, Wilder-Smith A, Bergeri I, Feikin DR, Arora RK, Patel MK, Subissi L. Protective effectiveness of previous SARS-CoV-2 infection and hybrid immunity against the omicron variant and severe disease: a systematic review and meta-regression. Lancet Infect Dis. 2023 Jan 18;23(5):556–67.

13. de La Vega MA, Polychronopoulou E, Xiii A, Ding Z, Chen T, Liu Q, Lan J, Nepveu-Traversy ME, Fausther-Bovendo H, Zaidan MF, Wong G, Sharma G, Kobinger GP. SARS-CoV-2 infection-induced immunity reduces rates of reinfection and hospitalization caused by the Delta or Omicron variants. Emerg Microbes Infect. 2023 Dec;12(1):e2169198.

14. Carazo S, Skowronski DM, Brisson M, Sauvageau C, Brousseau N, Gilca R, Ouakki M, Barkati S, Fafard J, Talbot D, Gilca V, Deceuninck G, Garenc C, Carignan A, De Wals P, De Serres G. Estimated Protection of Prior SARS-CoV-2 Infection Against Reinfection With the Omicron Variant Among Messenger RNA-Vaccinated and Nonvaccinated Individuals in Quebec, Canada. JAMA Netw Open. 2022 Oct 3;5(10):e2236670. [https://jamanetwork.com/journals/jamanetworkopen/fullarticle/2797311]

15. Carazo S, Skowronski DM, Brisson M, Sauvageau C, Brousseau N, Fafard J, et al. Prior infection-and/or vaccine-induced protection against Omicron BA.1, BA.2 and BA.4/BA.5-related hospitalisations in older adults: a test-negative case-control study in Quebec, Canada. Lancet Healthy Longev 2023 Aug;4(8):e409-e420.

16. Link-Gelles R, Ciesla AA, Roper LE, Scobie HM, Ali AR, Miller JD, Wiegand RE, Accorsi EK, Verani JR, Shang N, Derado G, Britton A, Smith ZR, Fleming-Dutra KE. Early Estimates of Bivalent mRNA Booster Dose Vaccine Effectiveness in Preventing Symptomatic SARS-CoV-2 Infection Attributable to Omicron BA.5- and XBB/XBB.1.5-Related Sublineages Among Immunocompetent Adults - Increasing Community Access to Testing Program, United States, December 2022-January 2023. MMWR Morb Mortal Wkly Rep. 2023 Feb 3;72(5):119-124.

17. Arbel R, Peretz A, Sergienko R, Friger M, Beckenstein T, Duskin-Bitan H, Yaron S, Hammerman A, Bilenko N, Netzer D. Effectiveness of a bivalent mRNA vaccine booster dose to prevent severe COVID-19 outcomes: a retrospective cohort study. Lancet Infect Dis. 2023 Apr 13:S1473-3099(23)00122-6.

18. Lin DY, Xu Y, Gu Y, Zeng D, Wheeler B, Young H, Sunny SK, Moore Z. Effectiveness of Bivalent Boosters against Severe Omicron Infection. N Engl J Med. 2023 Feb 23;388(8):764-766

19. Tenforde MW, Weber ZA, Natarajan K, Klein NP, Kharbanda AB, Stenehjem E, et al. Early Estimates of Bivalent mRNA Vaccine Effectiveness in Preventing COVID-19-Associated Emergency Department or Urgent Care Encounters and Hospitalizations Among Immunocompetent Adults - VISION Network, Nine States, September-November 2022. MMWR Morb Mortal Wkly Rep. 2023 Mar 17;71(53):1637-1646.

20. Surie D, DeCuir J, Zhu Y, Gaglani M, Ginde AA, Douin DJ, et al; IVY Network. Early Estimates of Bivalent mRNA Vaccine Effectiveness in Preventing COVID-19-Associated Hospitalization Among Immunocompetent Adults Aged ≥65 Years - IVY Network, 18 States, September 8-November 30, 2022. MMWR Morb Mortal Wkly Rep. 2022 Dec 30;71(5152):1625-1630.

21. Møller Kirsebom FCM, Andrews N, Stowe J, Ramsay M, Bernal JL. Duration of protection of ancestral-strain monovalent vaccines and effectiveness of bivalent BA.1 boosters against COVID-19 hospitalisation in England: a test-negative case control study. Lancet Infect Dis 2023 (in press). https://www.thelancet.com/journals/laninf/article/PIIS1473-3099(23)00365-1/fulltext

22. WHO. Vaccine efficacy, effectiveness and protection 2021. World Health Organization https://www.who.int/news-room/feature-stories/detail/vaccine-efficacy-effectiveness-and-protection (accessed 2 May, 2023).

23. Wang Q, Iketani S, Li Z, Liu L, Guo Y, Huang Y, Bowen AD, Liu M, Wang M, Yu J, Valdez R, Lauring AS, Sheng Z, Wang HH, Gordon A, Liu L, Ho DD. Alarming antibody evasion properties of rising SARS-CoV-2 BQ and XBB subvariants. Cell 2023;186(2):279-286.

24. Tan CY, Chiew CJ, Pang D, Lee VJ, Ong B, Wang LF, Ren EC, Lye DC, Tan KB. Effectiveness of bivalent mRNA vaccines against medically attended symptomatic SARS-CoV-2 infection and COVID-19-related hospital admission among SARS-CoV-2-naive and previously infected individuals: a retrospective cohort study. Lancet Infect Dis. 2023 Aug 2:S1473-3099(23)00373-0.

25. FDA. Future Vaccination Regimens Addressing COVID-19. Briefing document, Vaccines and Related Biological Products Advisory Committee Meeting, January 26, 2023. [https://www.fda.gov/media/164699/download]

26. COVID-19 Immunity Task Force. Seroprevalence in Canada. https://www.covid19immunitytaskforce.ca/seroprevalence-in-canada/

27. Tomic A, Skelly DT, Ogbe A, O’Connor D, Pace M, Adland E, et al. Divergent trajectories of antiviral memory after SARS-CoV-2 infection. Nat Commun. 2022 Mar 10;13(1):1251.

28. Boyton RJ, Altmann DM. The immunology of asymptomatic SARS-CoV-2 infection: what are the key questions? Nat Rev Immunol. 2021 Dec;21(12):762–768.

